# Finerenone Prescriptions in the US (2021-2024) by Physician Specialty: Analysis of Use and Potential in the CKM Space

**DOI:** 10.1101/2025.10.02.25337213

**Authors:** Yara Jelwan, Tianyu Cao, Zhiqi Yao, Semenawit Burka, John Erharbor, Phillip Berning, Omar Dzaye, Michael Blaha

**Affiliations:** Johns Hopkins Ciccarone Center for Cardiovascular Disease Prevention; Johns Hopkins Bloomberg School of Public Health Baltimore; University of Texas Southwestern Medical Center, Dallas, Texas, United States

## Abstract

**Introduction:** Finerenone, a non-steroidal mineralocorticoid receptor antagonist (MRA), has demonstrated greater receptor selectivity and fewer side effects compared to older mineralocorticoid receptor antagonists. Clinical trials support its efficacy in reducing kidney disease progression, cardiovascular events, and heart failure outcomes across various patient populations, however, anecdotally, the drug appears infrequently utilized in clinical practice.

**Methods:** A serial, cross-sectional analysis of IQVIA’s National Prescription Audit (NPA), which covers over 70% of U.S. outpatient prescription activity, was conducted from July 2021 to December 2024 to examine finerenone prescribing trends focusing on cardiologists and nephrologists. For added context, prescription rates of finerenone were compared to those of two other medications with related mechanisms or indications commonly used in Cardiovascular-Kidney-Metabolic (CKM) syndrome, spironolactone and empagliflozin. Prescribing trends were correlated with total search activity using Google Trends.

**Results:** We found that finerenone was prescribed at a rate of 30,180 prescriptions/month in December 2022, increasing to 45,420 in December 2023, and 60,756 prescriptions/month in December 2024. This modest increase correlated with changes in Google search activity, with mild inflections after the release of major clinical trial data. The ratio of total prescriptions from nephrologists (15.81 prescriptions per nephrologist) to cardiologists (0.70 prescriptions per cardiologist) in 2024 was approximately 23:1. The proportion of finerenone prescriptions to other CKM therapies remained low, with finerenone prescriptions numerically representing 3.0% of empagliflozin prescriptions and 2.6% of spironolactone prescriptions in 2024.

**Conclusion:** The adoption of finerenone has been modest, especially among cardiologists compared to nephrologists, and lags behind other CKM therapies. However, recent trends show an upward shift in its use that correlates with public interest related to clinical trial results.

## Introduction

Cardiovascular disease (CVD), chronic kidney disease (CKD), and metabolic conditions share common pathophysiological pathways and frequently coincide in the same population^1–2^. This overlap, known as Cardiovascular-Kidney-Metabolic (CKM) syndrome, has been recently defined by the American Heart Association as a systemic disorder involving interactions between metabolic risk factors, chronic kidney injury, and the cardiovascular system^3^. CKM has been recognized as a major determinant of premature morbidity and mortality; it affects individuals at risk for CVD, as well as those with existing CVD and is worsened by CKD and metabolic risk factors^4^.

Several targeted therapies have emerged, simultaneously addressing and improving those interconnected disease states in CKM syndrome^4^. Among these, mineralocorticoid receptor antagonists (MRAs) are a class of medications that have long been used to manage multiple components of that syndrome. Mineralocorticoid receptor (MR) activation in the distal nephron enhances sodium reabsorption and potassium excretion, leading to volume expansion, hypertension, and electrolyte imbalance. In non-epithelial tissues such as the heart and interstitium, inappropriate or excessive MR activation induces pro-inflammatory and pro-fibrotic gene expression, mediated by oxidative stress, transforming growth factor-β (TGF-β), and connective tissue growth factor (CTGF)^6–7^. These pathways promote tissue remodeling, fibrosis, and organ dysfunction, contributing to the progression of CKD and CVD^8^. By blocking the binding of aldosterone to the MR, MRAs reduce these pathological downstream effects, thus reducing fluid retention, lowering blood pressure, and protecting against organ damage, particularly in conditions like CKD and heart failure (HF)^5^.

Historically, steroidal MRAs such as spironolactone and eplerenone have been used in the management of HF and hypertension. More recently, finerenone, a non-steroidal MRA, has demonstrated greater selectivity and fewer side effects compared to older MRAs with multiple clinical trials supporting its use in various conditions across the CKM spectrum^5^. For example, FIDELIO-CKD trial demonstrated that finerenone significantly reduced the progression of kidney disease in patients with CKD and T2DM^9^. Shortly after, in 2021 the FIGARO-DKD trial showed that finerenone was associated with decreased cardiovascular events in a similar study population^10^. In 2024, the FINEARTS-HF trial showed that finerenone reduced the risk of the primary composite outcome of cardiovascular death and total HF events in patients with HF with Mid-Range Ejection Fraction (HFmrEF) and HF with preserved Ejection Fraction (HFpEF)^11^.

Finerenone has received varying levels of recommendation in both American and European clinical practice guidelines. The *American Kidney Disease Improving Global Outcomes (KDIGO) 2022 Clinical Practice Guideline for Diabetes Management in Chronic Kidney Disease* gives finerenone a Class 2A recommendation for use in patients with T2DM, an estimated glomerular filtration rate≥25 ml/min per 1.73 m², normal serum potassium concentration, and albuminuria (≥30 mg/g or ≥3 mg/mmol), despite the maximum tolerated dose of a renin-angiotensin system inhibitor (RASi)^12^. This contrasts with the Class 1A recommendation for sodium-glucose co-transporter 2 inhibitors (SGLT2i) for a similar patient population. In Europe, finerenone has been given a Class 1 recommendation in the *2023 European Society of Cardiology Guidelines for the management of cardiovascular disease in patients with diabetes*^13^, highlighting its role in reducing both cardiovascular and kidney failure risks when added to standard care.

However, despite its apparent efficacy and support from various societies, anecdotal reports suggest low utilization of finerenone by different physician specialties^14^, particularly cardiologists. This study aims to quantify recent trends in patterns of prescription of finerenone in the United States, with an emphasis on different specialties and a comparison to other therapies targeting the CKM system.

## Methods

We performed a serial, cross-sectional analysis of IQVIA’s National Prescription Audit (NPA). The NPA has been utilized in cardiovascular-related studies to analyze prescription trends and patterns in clinical practice^15–16–17^. The NPA represents and captures over 70% of all prescription activity in the United States, including Alaska and Hawaii, and covers all products, classes, and manufacturers^18^. The NPA then provides dispensing estimates based on a sample of ≈90% of all retail prescriptions dispensed in the United States, including 90% of retail pharmacies, 80% of nursing home pharmacies, and 70% of mail-service pharmacies. Data from IQVIA’s database was used to capture total finerenone prescribing trends across various medical specialties from the Association of American Medical Colleges (AAMC) and then specific trends for the cardiology and nephrology specialties.

Finerenone in this analysis is referred to as the total aggregate of all relevant products including the chemical name finerenone and brand name Kerendia.

To compare prescription rates between cardiologists, nephrologists and the total number of provider prescriptions, we graphed the total annual prescriptions of finerenone by each specialty from July 2021 to December 2024, in comparison to the total annual prescriptions by all providers.

To standardize the comparison per individual prescriber, we then divided the estimated yearly prescription volume of each specialty by the total number of cardiologists and nephrologists in the United States, respectively, obtaining the average annual prescriptions per physician for each specialty. We obtained the number of physicians in different specialties from the AAMC US Physician Workforce Data Report in 2023, which draws from the American Medical Association Physician Masterfile^19^. This approach allowed us to assess the most recent relative contribution of each specialty to finerenone prescribing.

We also directly compared finerenone prescription to that of spironolactone, the most used MRA, and empagliflozin, the first SGLT2i on the market. Both are older medications that are established in CKM syndrome with similar indications. Given that finerenone is a new drug on the market, we chose to compute the ratio of finerenone prescriptions to these other CKM therapies on a year-by-year basis to properly account for changing trends over time. For spironolactone and empagliflozin, national prescription data from 2019 and 2020 were included to provide baseline prescribing patterns before finerenone was introduced. Finally, we compared total prescriptions of finerenone, spironolactone and empagliflozin by provider specialty.

Online search data for the United States were retrieved for the period between July 2021 and December 2024 using the Google Health Trends Application Programming Interface (API), as described in previous studies^17^. Google Health Trends captures a sample of all public Google searches, including data from both signed-out and some signed-in users. While it excludes infrequent searches and duplicate queries from the same user within a short time frame, Google

Health Trends cannot entirely exclude the possibility that the same user may use multiple devices. Online search data were extracted from Google Health Trends as query fractions per 10 million searches, representing approximately 89% of all online searches for all individual drug brand names and chemical names.

### Statistical Analysis

We used descriptive statistics to perform our analyses. We evaluated over 197 million estimated prescriptions for finerenone, spironolactone, and empagliflozin, assessing monthly and yearly changes in prescription volumes. To compare relative use across the three drugs, we calculated the proportion of total prescriptions represented by each drug. We also examined annual prescriptions dispensed per physician within cardiology and nephrology specialties. These outcomes were visualized over time to identify notable trends and inflection points, which we then contextualized by overlaying key clinical trial publications and Food and Drug Administration (FDA) drug label expansions. All analyses were performed using R and Microsoft Excel.

### Funding

No extramural funding was used to support this work.

## Results

### Monthly prescription trends in the US and correlation with Google search activity

Between July 2021 and November 2024, the total monthly prescription rates of finerenone in the United States exhibited a consistent upward trend (Figure 1). Initially, prescription numbers started at 568 in June 2021 within the first year of the introduction of finerenone/Kerendia to the market. This increase continued through 2023 and 2024, reaching peak levels by the latest recorded data in December 2024. In parallel, online search interest for both “finerenone” and its brand name “Kerendia” followed a similar trajectory, with fluctuations corresponding to specific time points: the release of the 2022 KDIGO guidelines in November 2022 and the publication of the results of the FINEARTS-HF trial in September 2024 (Figure 1).

**Figure 1:**
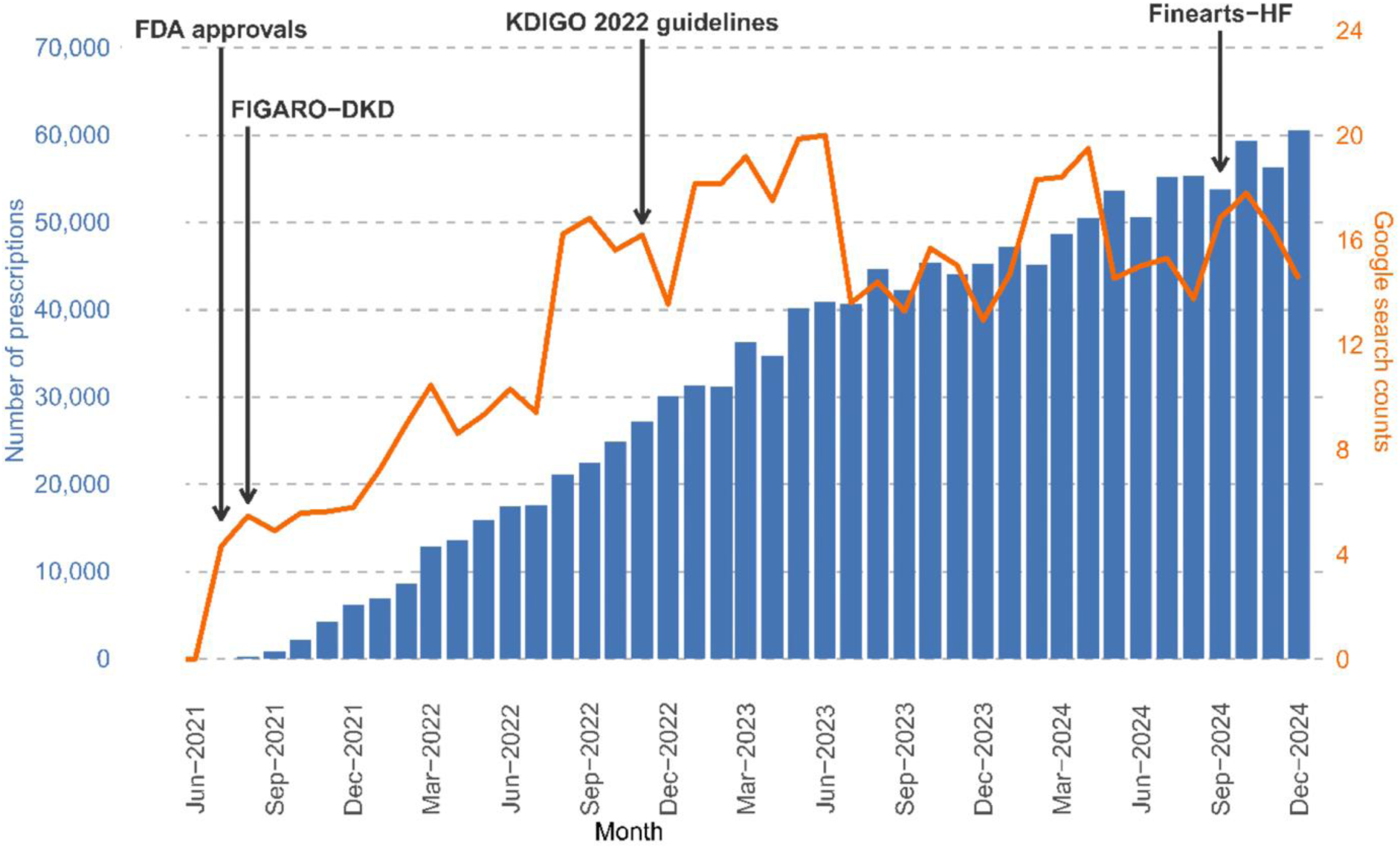
Monthly prescription trends for finerenone in the US across all specialties and correlation with Google search activity. Monthly prescription volumes (blue bars, left y-axis) are plotted alongside Google search counts for “finerenone” and “Kerendia” (orange line, right y-axis) from June 2021 through December 2024. Vertical arrows indicate the timing of key events, including FDA approval, publication of the FIGARO-DKD trial, release of the 2022 KDIGO guidelines, and the FINEARTS-HF trial.

### Annual prescription volumes overall and by specialties

From 2021 to 2024, the total number of annual finerenone prescriptions increased from 13,853 to 636,668 (Figure 2A). Between 2021 and 2024, annual prescriptions for finerenone increased across both cardiology and nephrology specialties (Figure 2 (A)). In 2021, nephrologists accounted for 4,269 prescriptions and cardiologists for 516. By 2024, nephrology prescriptions had risen to 195,372, while cardiology prescriptions reached 16,016. The proportion of total prescriptions attributed to nephrology was 30.8% (4,269/13,853) in 2021, 31.2% (68,326/218,799) in 2022, 30.5% (145,822/477,207) in 2023, and 30.7% (195,372/636,668) in 2024. The absolute number of prescriptions from cardiologists also increased each year, with the largest year-over-year change occurring between 2021 and 2022 (+6,412 prescriptions). Over the same period, the proportion of prescriptions attributed to cardiologists relative to the total amount of prescriptions was 3.7% (516/13,853) in 2021, 3.2% (6,928/218,799) in 2022, 2.6% (12,555/477,207) in 2023, and 2.5% (16,016/636,668) in 2024. While both specialties showed year-over-year increases in the absolute number of prescriptions, the relative proportion of total prescriptions remained higher and more stable for nephrologists compared to cardiologists over the study period.

**Figure 2.**
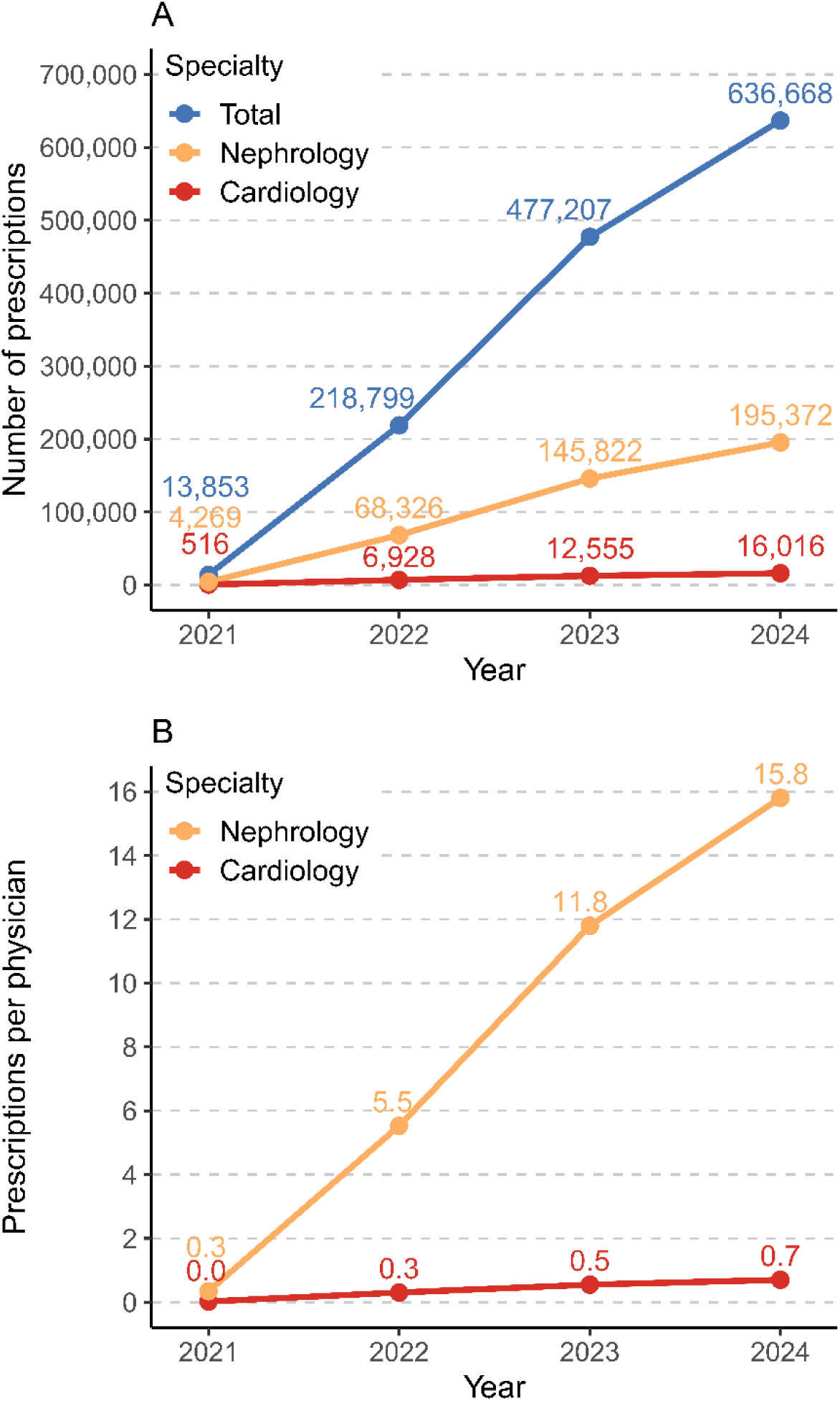
Annual prescriptions of finerenone by specialty in the United States. (A) Total annual finerenone prescriptions (blue) and prescriptions stratified by nephrology (orange) and cardiology (red) from 2021 through 2024. (B) Annual prescriptions per physician in nephrology and cardiology over the same period. Data demonstrate a greater absolute and per-physician uptake among nephrologists compared with cardiologists.

Using 2023 estimates of active physicians from the AAMC Physician Workforce Data Dashboard, the number of finerenone prescriptions per physician increased each year for both specialties. According to the 2023 U.S. Physician Workforce Data Dashboard, there were 22,843 physicians specializing in cardiovascular disease and 12,360 in nephrology in the United States and its territories. Among cardiologists, yearly prescriptions per physician were approximately 0.02 in 2021, 0.30 in 2022, 0.55 in 2023, and 0.70 in 2024 (Figure 2 (B)). Among nephrologists, the corresponding values were approximately 0.35 in 2021, 1.12 in 2022, 11.80 in 2023, and 15.81 in 2024.

### Comparison to the prescriptions of spironolactone and empagliflozin

Finerenone yearly prescriptions increased from 13,853 in 2021 to 636,668 in 2024, while spironolactone prescriptions rose from 19,458,298 to 24,328,627 over the same period (Table 1). The proportion of finerenone to spironolactone prescriptions increased from 0.07% in 2021 to 2.62% in 2024. Year-over-year, finerenone prescriptions increased by 33% between 2023 and 2024 (from 477,207 to 636,668), while spironolactone prescriptions increased by 7% (from 22,644,106 to 24,328,627). Over the full study period (2019–2024), a total of 1,346,527 prescriptions were written for finerenone and 123,416,916 for spironolactone.

**Table 1.** Annual prescription counts of mineralocorticoid receptor antagonist (MRA) drugs and comparative use of finerenone, 2019–2024. Values represent absolute prescription counts, with percentages in parentheses indicating year-over-year change. The rightmost column shows cumulative prescriptions across the six-year period and overall percent change from 2019 to 2024. The lower section reports finerenone prescriptions as a percentage of empagliflozin and spironolactone use.

|  | 2019 | 2020 | 2021 | 2022 | 2023 | 2024 | Total from<br>2019 to 2024 |
| --- | --- | --- | --- | --- | --- | --- | --- |
| **MRA drugs,**<br>**prescription**<br>**counts** |  |  |  |  |  |  |  |
| Finerenone | 0 | 0 | 13,853 | 218,799<br>(+1479%) | 477,207<br>(+118%) | 636,668<br>(+33%) | 1,346,527<br>(+9,620%) |
| Empagliflozin | 5,430,908 | 7,336,693<br>(+35%) | 9,537,827<br>(+30%) | 12,844,284<br>(+35%) | 16,785,904<br>(+31%) | 21,189,582<br>(+26%) | 73,125,198<br>(+290%) |
| Spironolactone | 17,656,902 | 18,560,062<br>(+5%) | 19,458,298<br>(+5%) | 20,768,921<br>(+7%) | 22,644,106<br>(+9%) | 24,328,627<br>(+7%) | 123,416,916<br>(+38%) |
| **Finerenone**<br>**prescriptions**<br>**relative to**<br>**other drugs** |  |  |  |  |  |  |  |
| vs<br>Empagliflozin | 0 | 0 | 0.15% | 1.70% | 2.84% | 3.00% | 1.84% |
| vs<br>Spironolactone | 0 | 0 | 0.07% | 1.05% | 2.11% | 2.62% | 1.09% |

Empagliflozin prescriptions increased from 9,537,827 in 2021 to 21,189,582 in 2024 (Table 1). In comparison, finerenone prescriptions rose from 13,853 to 636,668 during the same period. Finerenone prescription volume was equivalent to 0.15% (13,853/9,537,827) of empagliflozin prescriptions in 2021 and 3.00% (636,668/21,189,582) in 2024. Between 2023 and 2024, empagliflozin prescriptions increased by 26% (from 16,785,904 to 21,189,582), while finerenone prescriptions increased by 33%. Across the full study period, there were 73,125,198 prescriptions for empagliflozin and 1,346,527 for finerenone.

From January 2019 to July 2024, using a denominator of total prescriptions of finerenone + spironolactone + empagliflozin, the proportion of total prescriptions represented by spironolactone decreased steadily, from over 75% in early 2019 to just above 50% by mid-2024 (Figure 3). During the same period, empagliflozin’s share increased consistently, rising from approximately 25% in 2019 to nearly 45% by mid-2024. Finerenone, introduced in 2021, accounted for a small but gradually increasing share of prescriptions, reaching 1.5 % by 2024.

**Figure 3.**
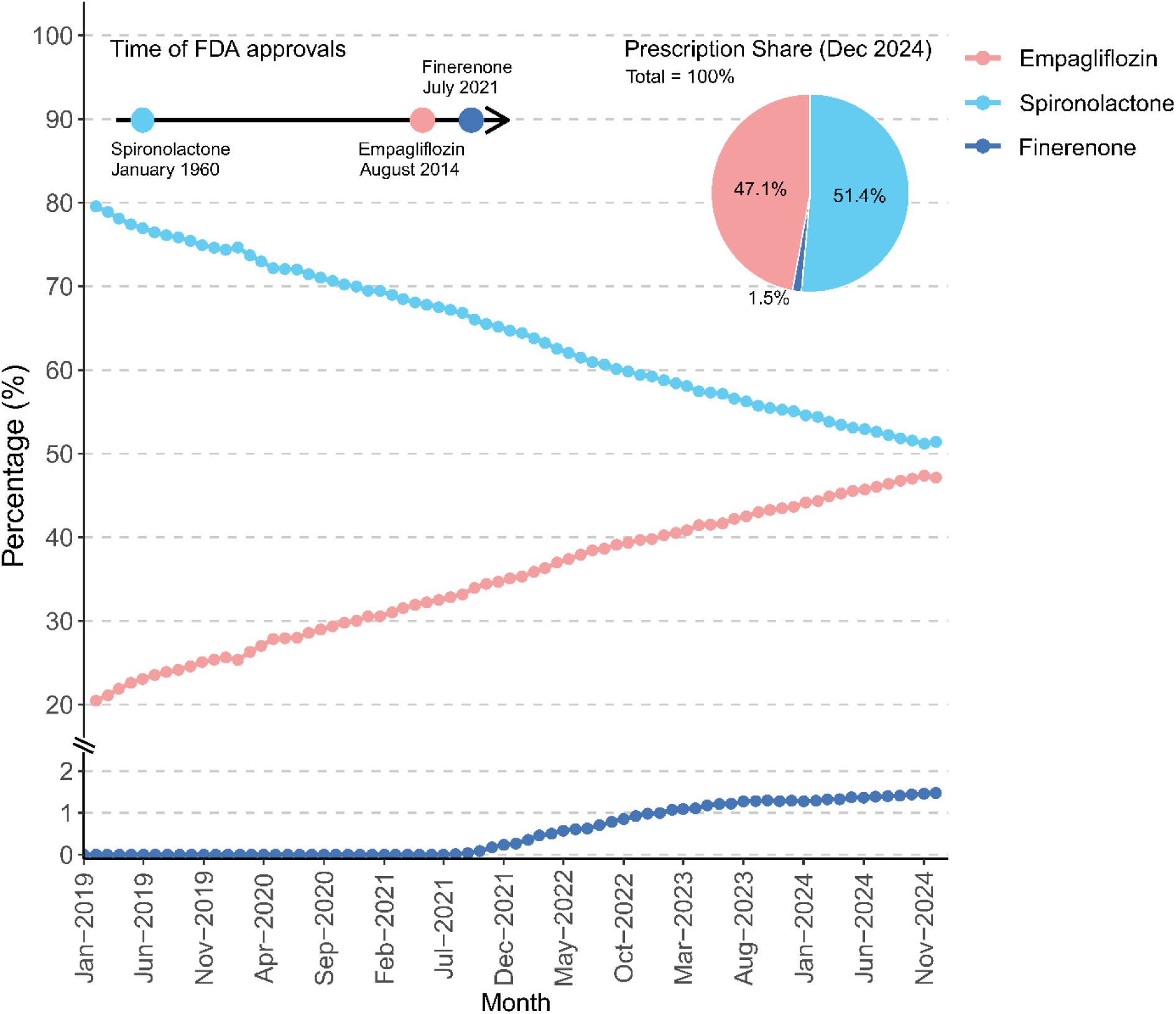
Relative prescription share of finerenone, spironolactone, and empagliflozin in the United States, 2019–2024. Monthly prescription proportions of spironolactone (blue), empagliflozin (red), and finerenone (dark blue) are shown from January 2019 through December 2024. Key FDA approval dates are indicated by arrows. The inset pie chart displays prescription share at the end of the study period (December 2024), with spironolactone accounting for 51.4%, empagliflozin 47.1%, and finerenone 1.5% of total prescriptions.

## Discussion

Despite strong evidence supporting the cardiorenal benefits of finerenone in patients with chronic kidney disease and T2DM, use of this therapy remains limited relative to other agents with overlapping indications. In this study, we analyzed a national audit of retail prescriptions to evaluate the uptake of finerenone by specialty since its market introduction in 2021. While total finerenone use increased substantially between 2021 and 2024, cardiologists accounted for a small proportion of overall prescriptions each year, with prescribing per cardiologist remaining markedly lower than that of nephrologists. In 2024, finerenone prescriptions per nephrologist were over 20 times higher than those per cardiologist. These findings parallel prior analyses of other cardiometabolic therapies, such as SGLT2i and glucagon-like peptide-1 (GLP-1) receptor agonists, which also demonstrated low initial rates of uptake by cardiologists despite strong clinical trial evidence and formal endorsement in guidelines^16^.

### Possible influence of key events on finerenone uptake

The observed trajectory of finerenone prescriptions over time may have been influenced by the timing of key clinical trial publications and regulatory approvals. Initial uptake followed the publication of FIDELIO-DKD in late 2020 and the subsequent FDA approval for treatment of CKD associated with T2DM in July 2021. A second inflection point occurred after the publication of the FIGARO-DKD trial in 2021, which expanded the evidence base to a broader population with earlier-stage CKD. Prescription volumes continued to rise steadily through 2023 and 2024, with an additional increase following the presentation of the FINEARTS-HF trial in late 2024, which evaluated finerenone in patients with HFpEF. The impact of the HFpEF indication will require a full evaluation in a future dedicated analysis using, for example, 2025 and 2026 data.

Despite these key milestones, search interest in finerenone, as assessed by Google Trends data, remained consistently lower than that of other medications in the same therapeutic space^16^.The volume of online searches for “finerenone” and the brand name “Kerendia” showed substantial peaks coinciding with trial publications or FDA approvals, demonstrating a modest increase in public interest following its market introduction. Monthly prescription data show a steady, linear growth pattern over time without abrupt surges, suggesting that dissemination of trial results and regulatory events may have contributed incrementally, but modestly, to prescriber behavior.

### Barriers to broader adoption

Several factors may contribute to the relatively modest uptake of finerenone, particularly among cardiologists. Although major trials have established its efficacy in reducing cardiorenal risk in patients with CKD and T2DM, U.S. clinical guidelines have not yet uniformly endorsed its use across specialties. The KDIGO guidelines provide a Class 2A recommendation for finerenone for CKD and T2DM indication^12^, but U.S.-based cardiology guidelines have only recently begun to reflect its potential role. In contrast, the European Society of Cardiology has assigned finerenone a Class 1A recommendation for CKD in patients with T2D^13^, reflecting a higher degree of endorsement compared to the US.

Familiarity and comfort with older mineralocorticoid receptor antagonists, specifically spironolactone, may also influence prescribing behaviors. Spironolactone remains widely used due to its longstanding clinical role, low cost, and broad guideline inclusion across heart failure and CKD populations. While finerenone offers a more selective pharmacologic profile and potentially lower risk of hyperkalemia, its newer status and lack of head-to-head trials with other MRAs may contribute to clinical inertia. Similarly, when compared to SGLT2i, recently introduced agents with broader indications, stronger Class 1 guideline recommendations, and earlier market entry, finerenone has seen lower adoption, despite overlapping therapeutic goals. In 2024, for example, finerenone prescriptions totaled less than 3% of those for empagliflozin.

Structural factors like cost and insurance coverage, also play a role. Like other newly approved therapies, finerenone may face coverage restrictions or require prior authorization, which can delay initiation^20^. Additionally, concerns about hyperkalemia and the need for ongoing laboratory monitoring may reduce enthusiasm among clinicians or patients, even when risks are relatively low^21^. Another factor for decreased use of finerenone might be due to the previous lack of data for use in conjunction with SGLT2 inhibitors.

Moreover, a multi-pillar approach to CKM had not been established until only recently. In a CKM space increasingly crowded with effective options for therapy, physicians appear to have prioritized agents they were already familiar with. New guidelines emphasizing a multi-pillar approach to both CKD and HFpEF will likely influence future finerenone prescribing.

Together, these factors highlight a persistent implementation gap between evidence generation and clinical practice. Targeted efforts to address specialty-specific barriers, through education, updated guidelines, and coordinated care models, may be necessary to fully realize the benefits of finerenone in appropriate patient populations.

There are a few limitations to this study. Foremost, our data runs only through 2024, so it should not be interpreted as a thorough evaluation of the effects of the new HFpEF indication. Our data reflect dispensed prescriptions and therefore cannot capture attempts to prescribe finerenone that were denied by insurance or did not meet pre-authorization requirements. Advanced practice providers remain difficult to categorize using these datasets, as the setting of their practice (primary care vs. cardiology vs. nephrology for example) are not reflected in the dataset.

## Conclusion

In conclusion, while finerenone has shown great potential in the CKM space, its adoption has been slow despite its inclusion in guidelines, and this appears to be reflected in Google search data. The rate of adoption is particularly slower among cardiologists compared to nephrologists its use remains limited when compared to other CKM therapies like spironolactone and SGLT2 inhibitors. To improve evidence-based integration into clinical practice, more education and updated guidelines may be needed to better inform healthcare providers on finerenone’s potential efficacy and appropriate use. Future analyses will be needed using future prescribing data to study the effect of the new HFpEF indication.

## Data Availability

All data referred to in this manuscript are available from the corresponding author upon reasonable request.

## Acknowledgments

The authors thank all colleagues who provided input and support.

